# Insights into Hypertension: Microbial Load, Salivary Cytokine Levels and Salivary Flow in Edentulous Patients With Complete Dentures: a Case-Control Study

**DOI:** 10.1101/2024.05.15.24307456

**Authors:** Pillar Gonçalves Pizziolo, Lorena Mosconi Clemente, Aline Barbosa Ribeiro, Viviane de Cássia Oliveira, Ana Paula Macedo, Helio Cesar Salgado, Rubens Fazan, Evandro Watanabe, Cláudia Helena Lovato da Silva, Adriana Barbosa Ribeiro

**Author notes:** CORRESPONDING AUTHOR, Cláudia Helena Lovato da Silva, Department of Dental Materials and Prosthesis, Ribeirão Preto School of Dentistry, University of São Paulo, Café Avenue S/N, Ribeirão Preto 14040-904, SP, Brazil.

## Abstract

**BACKGROUND:** Tooth loss has been established as a correlate with an increased risk of cardiovascular disease (CVD) and hypertension. However, the influence of complete denture usage on hypertension remains inadequately explored in scientific literature.

**METHODS:** This case-control study evaluated the microbial load of the biofilm of dentures and the palate, salivary levels of cytokines (IL-2, IL-4, IL-6, IL-10, TNF-α, IFN-γ, and IL-17) and Salivary Flow (SF) of healthy (CG), controlled hypertensives (G1), underreported hypertensives (G2) and uncontrolled hypertensives (G3). The sample was characterized by sociodemographic data, clinical information, systolic (SBP), and Diastolic Blood Pressure (DBP). The microbial load of *Candida* spp., *Staphylococcus* spp., enterobacteria, and mutans group streptococci was evaluated by quantifying colony-forming unit (CFU), SF of unstimulated saliva by mL per minute, and salivary cytokines by flow cytometry.

**RESULTS:** 80 patients were evaluated (66±7.2 years). The time of edentulism was longer in G3 and positively associated with SBP. The CFU of mutans group streptococci on the denture was higher in G3 and showed a negative association with smoking habit, and this had a positive association with salivary cytokines (IL-4, IL-2, IL-17, IFN-γ), diabetes, and CVD. Patients in group G3, who only use upper dentures, had significantly higher SBP (p=0.024) and levels of IL-2 (p=0.024).

**CONCLUSIONS:** Time of edentulism may play a role in hypertension. Smoking habits modulated microbiota and interleukins, especially in diabetic and CVD patients. Furthermore, non-functional dentures had an association with hypertensive patients not controlled by medication, reflected in an increase in SBP and IL-2.

## INTRODUCTION

Previous studies suggest that inadequate oral health may be related to the increased risk of mortality, cardiovascular and respiratory diseases,^1,2^ and it is worth highlighting has been included as an unconventional risk factor for cardiovascular diseases.^3–5^ Furthermore, several studies have shown an association between tooth loss and elevated blood pressure,^6–11^ as well as partial and total edentulism with stroke.^12–14^

Recent research findings emphasize that the demographic profile of completely edentulous patients often consists of elderly individuals.^15–17^ Advanced age is recognized as a factor in the manifestation of numerous diseases, spanning atherosclerosis, diabetes mellitus, Alzheimer’s disease, rheumatoid arthritis, neoplastic disorders, and the overarching process of senescence.^18^ Furthermore, the aging process is associated with an upsurge in inflammatory biomarkers and increased colonization of pathogenic microorganisms within the saliva,^19^ predisposing these individuals to the worsening of systemic diseases.^18^

In this context, denture wearers constitute a demographic susceptible to potential alterations in the oral microbiome composition, marked by reduced microbial diversity and heightened prevalence of pathogenic bacteria and fungi.^5,19,20^ Moreover, individuals utilizing complete dentures frequently encounter denture stomatitis (DS), a chronic inflammatory condition linked to increased salivary levels of IL-6 and the proliferation of *Candida albicans*.^16^ DS has been associated with endothelial dysfunction,^21^ and its management has been shown to ameliorate endothelial function,^22^ systolic blood pressure (SBP), and the expression of cytokines IL-6, IL-2, IL-10, and IFNγ among elderly edentulous patients.^15^

The association between oral infections and cardiovascular disease (CVD) is attributed to knowledge of biological mechanisms, including transient bacteremia, translocation of oral bacterial pathogens, and elevated levels of circulating inflammatory biomarkers.^21, 23, 24^ Another proposed theory suggests a link between oral health status and CVD risk, positing that the diminished chewing efficiency often observed in complete denture wearers may significantly impact oral functions.^2^ This compromised function can potentially lead to nutritional deficiencies, obesity, elevated cholesterol levels, and chronic systemic inflammation, thereby increasing the risk of developing CVD.^24–27^

Numerous theories have proposed a link between oral health and CVD, even so, a gap exists in comprehending the oral and systemic characteristics of normotensive and hypertensive individuals who wear complete dentures. This study aims to evaluate the microbial load of denture and palate biofilms, salivary flow, and levels of inflammatory cytokines in normotensive and hypertensive edentulous patients wearing complete dentures. The null hypothesis posits that there are no significant differences in microbial load, salivary flow rates, and inflammatory cytokine levels between the experimental and control groups.

## METHODS

### Study Design

This is a retrospective observational case-control clinical study, with a sample composed of fully edentulous individuals who are users of conventional complete dentures and have good general health conditions (control group), and those afflicted with hypertension (cases group). The study design will be reported following the STROBE statement for observational clinical studies. All information related to health conditions, blood pressure, medications used, and history of deleterious habits such as smoking and alcohol consumption, was obtained for each group (cases and control) to enable an analysis of group parity trends.

### Study Population

This study was approved by the Human Research Ethics Committee of the Ribeirão Preto School of Dentistry, University of São Paulo (FORP/USP) (CAAE: 93712418.1.0000.5419). All participants provided written informed consent and were recruited between March 2022 and October 2023. The study included edentulous individuals using conventional complete dentures who received treatment in the complete dentures discipline at FORP/USP.

The inclusion criteria for this study comprised individuals of both sexes, either in good general health (control group) or diagnosed with hypertension (case group), who had been utilizing either both dentures or only the upper complete denture in satisfactory condition for at least 1 year. The dentures were constructed from heat-polymerized acrylic resin and acrylic teeth. Exclusion criteria included individuals with pacemakers, atrial fibrillation/flutter, complete dentures exhibiting adaptation issues, relining, repairs, or fractures. Additionally, individuals who were immunosuppressed, undergoing chemotherapy or radiotherapy, or had taken antibiotics, anti-inflammatories, or antifungal agents within the preceding four weeks were excluded. Individuals presenting with oral mucosal lesions such as denture-induced fibrous hyperplasia, papillomas, or traumatic ulcerations associated with denture bases were also excluded from the study. According to these criteria, patients were divided into four groups:

The sample size was estimated based on systolic blood pressure. A previous study provided values for the expected standard deviations,^15^ considering a 95% (bilateral) confidence interval and a power of 80%. These parameters required at least 10 participants per group (power of 80%. α = 0.05; β = 0.20).

Control Group (CG) - Healthy: Completely edentulous individuals wearing conventional complete dentures, normotensive not taking antihypertensive medications, with systolic blood pressure below 140 mmHg and diastolic blood pressure below 90 mmHg measured during consultation.

Case Group 1 (G1) - Controlled hypertensives: Completely edentulous patients wearing conventional complete dentures, undergoing treatment with at least one antihypertensive medication (including diuretics), with systolic blood pressure below 140 mmHg and diastolic blood pressure below 90 mmHg measured during consultation.

Case Group 2 (G2) - Underreported hypertensives: Completely edentulous patients wearing conventional complete dentures, not taking antihypertensive medications, not yet diagnosed with hypertension, with systolic blood pressure surpassing 140 mmHg and diastolic blood pressure surpassing 90 mmHg as measured during consultation.

Case Group 3 (G3) - Uncontrolled hypertensives: Completely edentulous patients wearing conventional complete dentures, using at least one antihypertensive medication (including diuretics), with systolic blood pressure surpassing 140 mmHg and diastolic blood pressure surpassing 90 mmHg as measured during consultation.

Individuals with systolic blood pressure (SBP) exceeding 140 mmHg or diastolic blood pressure exceeding 90 mmHg, or those taking antihypertensive medication as per the Brazilian Arterial Hypertension Guideline^30^ were classified as hypertensive.

### Data Collection

Sociodemographic variables, behavioral information related to hygiene habits, nighttime use of dentures, smoking habits, and cleaning frequency, as well as information on oral and systemic health, were collected through specific questionnaires. Clinical information such as the presence of biofilm, edentulism time, Kapur index,^28^ and classification of denture-related stomatitis (Newton’s Classification modified by Kabawat)^29^ was evaluated by two researchers (A.B.R.; C.H.L.) were previously calibrated with a Kappa value of 0.878.

To minimize memory bias regarding self-reported hypertension diagnoses, blood pressure measurements were utilized to categorize patients into the control and case groups. The oscillatory sphygmomanometer method was employed, where three indirect blood pressure readings were obtained using an automated device (HEM7130, Omron Healthcare Brazil, Brazil, São Paulo, SP), with an interval of at least five minutes between measurements by the same researcher (P.G.P). Systolic and diastolic blood pressure values in mmHg were recorded, and the average of the three values obtained was calculated.

To analyze the microbial load, the same blind researcher (V.C.O.) placed the upper denture in Petri dishes, in an aseptic area, to desorb the biofilm with a sterilized brush (Tek, Bristles soft Johnson & Johnson do Brasil Indústria e Comércio de Produtos para Saúde Ltda., S. J. dos Campos, SP, Brazil) and buffered saline solution (PBS- phosphate buffered saline).^31,32^ To collect samples from the palate, another researcher (P.G.P.) rubbed a sterile cytology brush on the palatal mucosa regions.^29^ Then, the active tip was sectioned and stored in a sterile microtube containing 1.5 mL of PBS solution. The solutions were agitated using a vortex and then subjected to serial dilution from 10⁻¹ to 10⁻^3^ and seeded in Petri dishes with a specific culture medium for the growth of *Candida* spp. (CHROMagarTM Candida, Becton Dickinson, Paris, France), *Staphylococcus* spp. (Mannitol salt Agar, Kasvi Imp. e Dist. de Prod. para Laboratórios Ltda., Curitiba, Brazil), mutans group streptococci (SB20 Modified Agar)^33^ and enterobacteria (MacConkey Agar, Himedia Laboratories PVI Ltd., Mumbai, India), The plates were incubated in a microbiological oven at 37°C for 48 hours and mutans group streptococci were cultivated in an environment in an anaerobic jar. After the incubation period, the researchers (P.G.P., L.M.C., A.B.R.) performed the colony-forming unit (CFU) count to quantify the microbial load using the formula CFU/mL = n° de count x 10ᵑ/q.^31^

The salivary flow was calculated by the volume, in mL, per minute of unstimulated saliva collected for 10 minutes, between 9 and 11 am, using the spitting method. Quantification of the concentration of salivary levels of cytokines IL-2, IL-4, IL-6, IL-10, TNF-α, IFN-γ and IL-17 was carried out by a blind researcher (A.B.R.2) using the commercial kit of BD™ Cytometric Bead Array (CBA) Human Th1/Th2/Th17 (RRID: AB_2869353) Cytokine (BD; San Jose, CA, USA), according to the manufacturer’s instructions.

### Data analysis

Qualitative data (sociodemographic characteristics, behavioral characteristics, presence of biofilm, Kapur index, presence of burning mouth, xerostomia, and degree of DS) were shown in a cross-reference table (contingency tables) and analyzed by the exact test Fisher’s or likelihood ratio. The quantitative variables (age, cleaning frequency, edentulism time, age of dentures in use, blood pressure, medications, microbial load, salivary flow, and cytokines) were subjected to the normality test (Shapiro-Wilk) and homogeneity of variance test. (Levene). Once these criteria were met, the ANOVA tests and Tukey post-test with Bonferroni adjustment were used (age and blood pressure). Results that did not meet the normality and homoscedasticity criteria were analyzed with the Kruskal-Wallis test and Dunn and Mann-Whitney post-tests (DS degree, microbial load, salivary flow, and cytokines). Correlation analysis was performed using the Spearman test. All tests were performed with the SPSS 21.0 statistical software (SPSS Inc., Chicago, IL, USA) by a blind researcher (A. P. M.), considering a significance level of 5%.

## RESULTS

The initial sample comprised 114 participants evaluated for eligibility, of whom 80 met the criteria and were included in the final sample, allocated across four groups (CG, G1, G2, and G3) with n=20 (Figure S1). The sample was mostly composed of elderly people (66.7±7 years), between 61 and 70 years old (55.0%), and no statistically significant difference between the groups (*P*=0.170). Regardless of group, elderly patients are positively associated with systolic blood pressure (*P*=0.028; r=0.245) and hypertension (*P*=0.038; r=0.232), as shown in Figure 1. No significant differences were found among the four groups for other sociodemographic characteristics (Table S1).

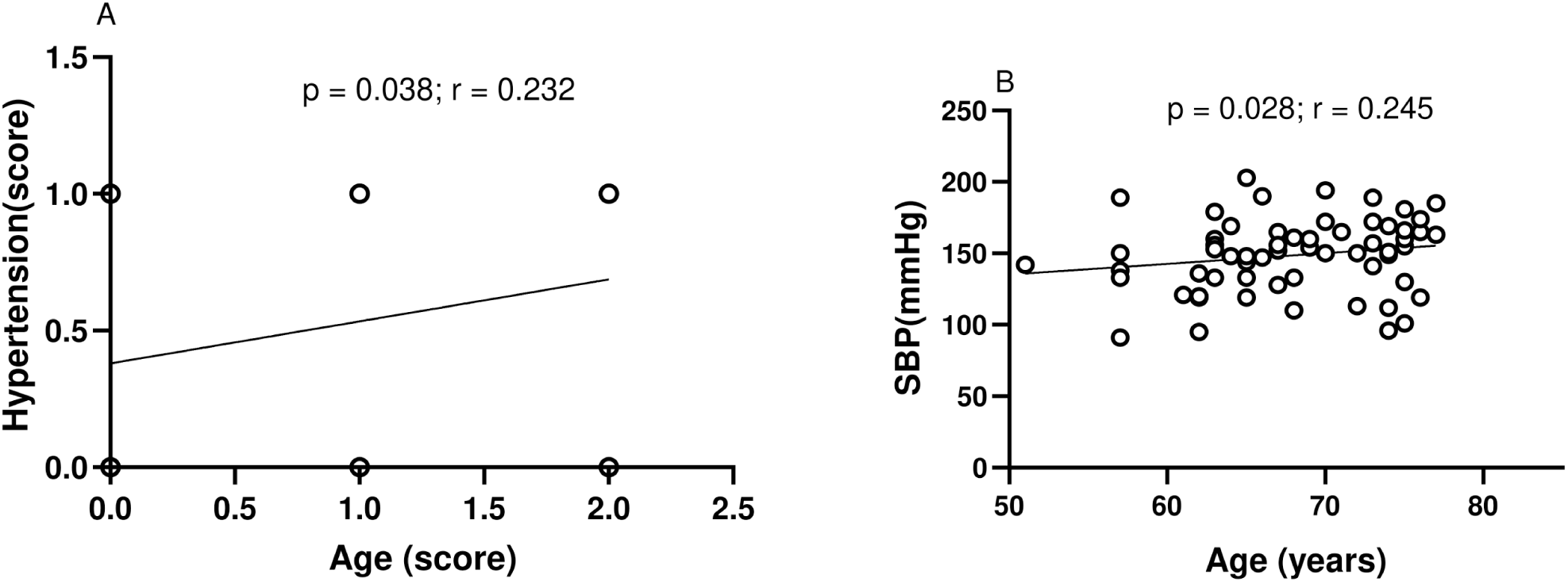

There was no statistical difference regarding behavioral characteristics between the groups, such as removing dentures at night (upper denture *P*=0.344; lower denture *P*=0.090), alcohol consumption (*P*=0.724), and frequency of denture hygiene (*P*=0.378). However, when observing smoking habits, G1 had a higher frequency of smokers (55.0%), and ex-smokers (38.9%) compared to the other groups (*P*=0.042) (Table 1). Furthermore, smoking habits were not associated with systolic blood pressure (*P*=0.069; r=-0.262) and diastolic blood pressure (*P*=0.365, r=-0.132). However, there was an association between the number of cigarettes consumed daily and CFU of mutans group streptococci in the dentures (*P*=0.017; r=-0.338), salivary levels of IL-4 (*P*=0.026; r=0.318), IL- 2 (*P*=0.007; r=0.381), IL-17 (*P*=0.003; r=0.420) and IFN-γ (*P*=0.033; r=0.305), diabetic patients (*P*=0.033; r=0.304) and with CVD (*P*=0.048; r=0.284). *Staphylococcus spp.* in the dentures was associated with the patients with reported hypercholesterolemia (*P*=0.034; r=0.237), as shown in Table 2.

**Table 1.** Behavioral characteristics [n (%) or Mean (SD); Median (CI)] of study participants.

| Outcomes | Groups |  |  |  | P-value |
| --- | --- | --- | --- | --- | --- |
|  | CG | G1 | G2 | G3 |  |
| Upper denture removal at night |  |  |  |  |  |
| Always | 10 (52.6%) | 6 (30.0%) | 12 (60.0%) | 10 (50.0%) | 0.344* |
| Never | 1 (5.3%) | 1 (5.0%) | 0 (0.0%) | 2 (10.0%) |  |
| Sometimes | 8 (42.1%) | 13 (65.0%) | 8 (40.0%) | 8 (40.0%) |  |
| Lower denture removal at night |  |  |  |  |  |
| Always | 4 (26.7%) | 5 (35.7%) | 9 (52.9%) | 8 (61.5%) | 0.090* |
| Never | 3 (20.0%) | 0 (0.0%) | 0 (0.0%) | 1 (7.7%) |  |
| Sometimes | 8 (53.3%) | 9 (64.3%) | 8 (47.1%) | 4 (30.8%) |  |
| Smoking |  |  |  |  |  |
| Never | 9 (45.0%) | 1 (5.6%) | 9 (45.5%) | 11 (55.0%) | 0.042* |
| Past | 6 (30.0%) | 7 (38.9%) | 7 (35.5%) | 3 (30.0%) |  |
| Current | 5 (25.0%) | 10 (55.5%) | 4 (20.0%) | 3 (15.5%) |  |
| Number of cigarettes per day |  |  |  |  |  |
| Mean (SD) | 15.33 (12.69) | 21.50 (12.14) | 17.92 (10.55) | 12.00 (8.19) | 0.440** |
| Median | 12.00 | 20.00 | 20.00 | 12.00 |  |
| (CI) | (7.27;23.39) | (15.03;27.97) | (11.21;24.62) | (5.71;18.29) |  |
| Alcohol consumption |  |  |  |  |  |
| Never | 10 (50.0%) | 4 (20.0%) | 11 (57.9%) | 7 (35.5%) | 0.724* |
| Sometimes | 8 (40.0%) | 14 (70.0%) | 8 (42.2%) | 12 (60.0%) |  |
| Several times | 1 (5.0%) | 2 (10.0%) | 0 (0.0%) | 1 (5.0%) |  |
| Number of doses at a time |  |  |  |  |  |
| 1 to 2 | 2 (22.2%) | 9(56.2%) | 4 (66.7%) | 10 (76.9%) | 0.194* |
| 3 to 4 | 4 (44.4%) | 4 (25.0%) | 1 (16.7%) | 2 (15.4%) |  |
| 5 to 6 | 3 (33.3%) | 1 (6.2%) | 1 (16.7%) | 1 (7.7%) |  |
| More than 7 | 0 (0.0%) | 2 (12.5%) | 0 (0.0%) | 0 (0.0%) |  |

\* Fisher's exact test. \*\* Spearman test. SD=Standard deviation. CI=Confidence interval. Values in bold indicate significant statistical differences, considering ( $P$ -value<0.05).

**Table 2.** Correlation of Colony Forming Units in Log10 (CFULog10) of *Staphylococcus* spp. and mutans group streptococci in dentures and smoking habits with cardiovascular diseases, hypertension, diabetes, cholesterol, and use of diuretics drugs.

|  | Cardiovascular Disease | Hypertension | Smoking habit | Diabetes | Cholesterol | Diuretics |
| --- | --- | --- | --- | --- | --- | --- |
| *Staphylococcus* spp (CFU) | $P=0.981$<br>$r=0.003$ | $P=0.597$<br>$r=0.60$ | $P=0.933$<br>$r=-0.012$ | $P=0.912$<br>$r=0.013$ | **$P=0.034$**<br>$r=0.237$ | $P=0.766$<br>$r=0.034$ |
| mutans group streptococci | **$P=0.006$**<br>$r=-0.305$ | $P=0.339$<br>$r=0.108$ | **$P=0.017$**<br>$r=-0.338$ | $P=0.474$<br>$r=-0.081$ | $P=0.205$<br>$r=0.143$ | **$P=0.005$**<br>$r=-0.312$ |
| Smoking habit<br>Number of<br>cigarettes/days | **$P=0.048$**<br>$r=0.284$ | **$P=0.002$**<br>$r=-0.339$ | -<br>- | **$P=0.033$**<br>$r=0.284$ | $P=0.223$<br>$r=-0.177$ | **$P=0.015$**<br>$r=0.344$ |

Spearman test. Values in bold indicate significant statistical differences, considering ( $P$ -value<0.05).

Regarding oral clinical characteristics, there was a notable variance in the duration of edentulism across groups (*P*=0.031). Group G3 exhibited the highest median duration (40.00 years), followed by groups G2 (32.50 years), CG (26.50 years), and G1 (22.50 years) (Table 3). Moreover, the duration of edentulism exhibited a significant correlation with systolic blood pressure (*P*=0.012; r=0.281) (Figure 2A). Analysis of dentures quality and rehabilitation functionality revealed that patients who lacked functional rehabilitation, i.e., those with only upper complete dentures, demonstrated elevated systolic blood pressure in G3, with intermediate values observed in G1 (Figure 2B) (*P*=0.024). Additionally, levels of IL-2 cytokines were notably higher in this group, particularly in G3 (Figure 2C). Other evaluated oral clinical characteristics did not exhibit statistically significant differences (Table 3).

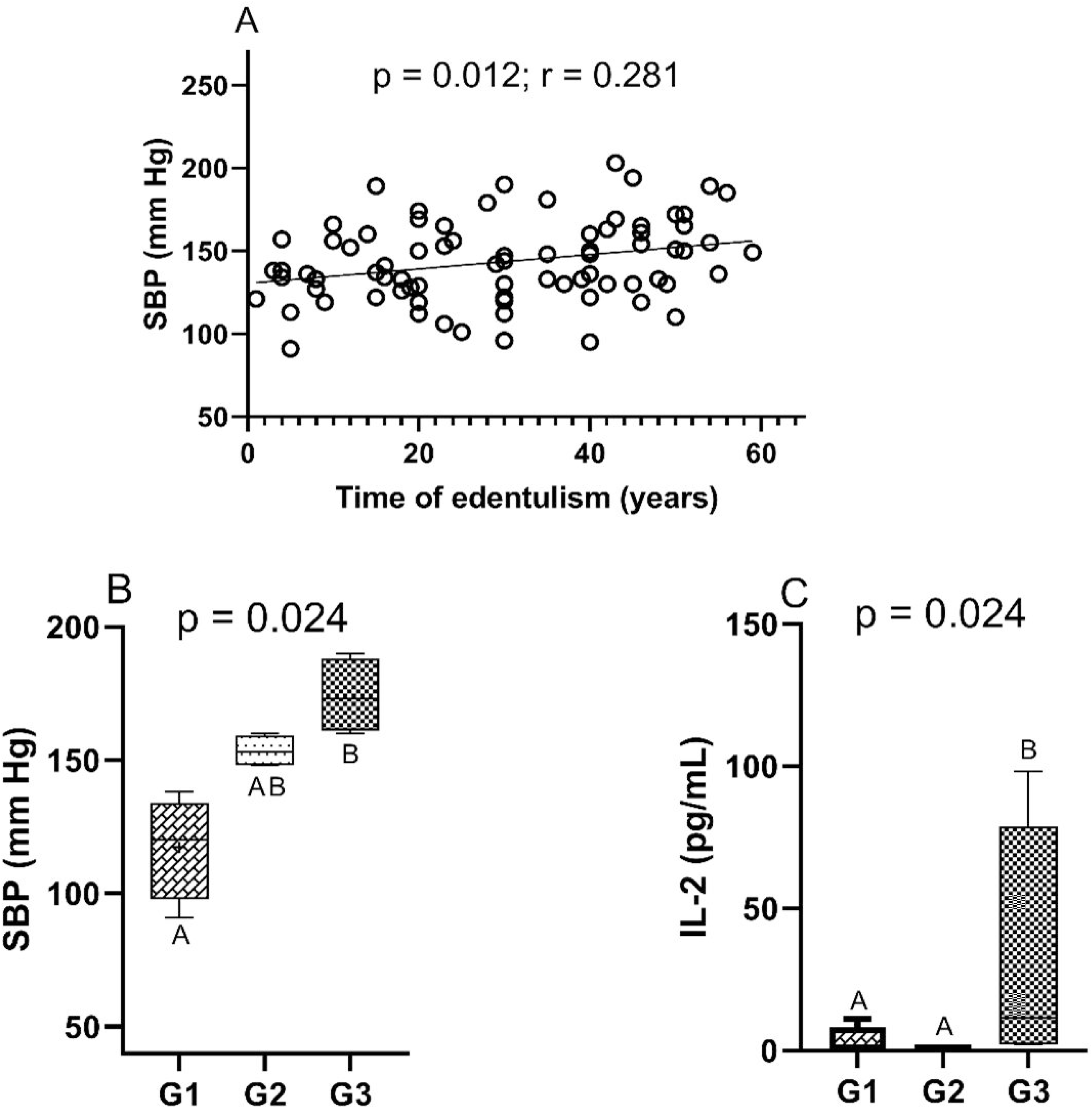

**Table 3.** Dental clinical characteristics [Mean (SD); Median (CI)] of study participants.

| Outcomes |  | Groups |  |  |  | P-value |
| --- | --- | --- | --- | --- | --- | --- |
|  |  | CG | G1 | G2 | G3 |  |
| Presence of Biofilm | Mean | 1.50 | 1.35 | 2.25 | 1.60 | 0.346** |
|  | (SD) | 1.15 | 1.18 | 0.93 | 1.27 |  |
|  | Median | 1.50<sup>a</sup> | 1.00<sup>a</sup> | 2.00<sup>a</sup> | 1.00<sup>a</sup> |  |
|  | (CI) | (0.96; 2.04) | (0.80; 1.90) | (1.71; 2.59) | (1.00; 2.20) |  |
| Time of edentulism | Mean | 26.10 | 24.35 | 34.70 | 34.05 | 0.031** |
|  | (SD) | 15.53 | 15.90 | 14.29 | 16.19 |  |
|  | Median | 26.50<sup>b</sup> | 22.50<sup>ab</sup> | 32.50<sup>b</sup> | 40.00<sup>b</sup> |  |
|  | (CI) | (18.83; 33.37) | (16.91; 31.79) | (28.01; 41.39) | (26.47; 41.30) |  |
| Age of upper dentures | Mean | 11.65 | 10.00 | 12.15 | 10.65 | 0.570** |
|  | (SD) | 10.51 | 9.49 | 13.95 | 11.97 |  |
|  | Median | 9.00<sup>a</sup> | 8.00<sup>a</sup> | 8.00<sup>a</sup> | 6.00<sup>a</sup> |  |
|  | (CI) | (6.73; 16.57) | (5.56; 14.44) | (5.62; 18.68) | (5.05; 16.25) |  |
| Age of lower dentures | Mean | 10.83 | 9.38 | 8.19 | 9.81 | 0.079** |
|  | (SD) | 10.71 | 7.93 | 6.87 | 10.62 |  |
|  | Median | 7.50<sup>a</sup> | 8.00<sup>a</sup> | 7.00<sup>a</sup> | 6.00<sup>a</sup> |  |
|  | (CI) | (5.51; 16.16) | (4.59; 14.18) | (4.53; 11.85) | (4.16; 15.47) |  |
| Denture Stomatitis Degrees |  |  |  |  |  |  |
| Yes |  | 12 (60.0%) | 11 (55.0%) | 11 (55.0%) | 10 (50.0%) | 0.756* |
| IA |  | 2 (4.5%) | 1 (2.27%) | 3 (6.8%) | 2 (4.5%) |  |
| IB |  | 3 (6.8%) | 3 (6.8%) | 3 (6.8%) | 2 (4.5%) |  |
| II |  | 5 (11.3%) | 3 (6.8%) | 2 (4.5%) | 3 (6.8%) |  |
| III |  | 2 (4.5%) | 4 (9.0%) | 3 (6.8%) | 3 (6.8%) |  |
| No |  | 8 (40.0%) | 9 (45.0%) | 9 (45.0%) | 10 (50.0%) |  |
| Kapur Index |  |  |  |  |  |  |
| Poor |  | 11 (50.0%) | 5 (25.5%) | 6 (30.0%) | 6 (30.0%) | 0.457* |
| Satisfactory |  | 5 (25.5%) | 6 (30.0%) | 8 (40.0%) | 7 (35.0%) |  |
| Good |  | 4 (20.0%) | 9 (45.5%) | 6 (30.0%) | 7 (35.5%) |  |

\* Fisher's exact test, \*\*Spearman test. Values in bold indicate significant statistical differences, considering ( $P\text{-value}<0.05$ ). SD=Standard deviation. CI=Confidence interval. DS= Denture stomatitis. IA: petechiae in normal palatal tissue; IB: localized area of inflammation of the denture-bearing area; II: generalized area of inflammation of the denture-bearing area; III: hyperplastic palatal surface with inflammations of the denture-bearing area. Different letters indicate statistical differences.

Concerning salivary parameters, patients in the CG reported a greater sensation of xerostomia (*P*=0.011). However, no discernible difference in salivary flow was observed among the groups (*P*=0.362) (Table S2). Additionally, male patients, irrespective of group allocation, exhibited lower salivary flow than females (*P*=0.008). A correlation was found between salivary flow and xerostomia, independent of group assignment, indicating that higher salivary flow corresponded to a reduced sensation of dry mouth (*P*=0.042; r=-0.228). Examination of the relationship between salivary flow and denture stomatitis (DS) degree revealed that patients with lower salivary flow tended to have a higher DS degree (*P*=0.046, r=-0.224). Although mean salivary cytokine levels were notably higher in G2, no statistically significant difference was observed between the groups (Table S2). However, individuals with more severe DS degrees exhibited elevated levels of TNF-α (*P*=0.010).

When assessing the distribution of DS within the study groups, it was found that 55% of the sample had the disease, with 9 (20.4%) classified as IA degree, 11 (25%) as IB, 13 (29.5%) as II, and 12 (27.2%) as III, demonstrating no statistically significant difference between the groups (*P*=0.756) (Table 3). However, upon investigating the association between the degree of DS and microbial load, irrespective of group allocation, individuals with more severe DS degrees exhibited higher colony-forming unit (CFU) counts for *Candida* spp. on the palate (*P*=0.030) and on the denture (*P*=0.020), *Candida albicans* on the denture (*P*=0.040), *Candida tropicalis* on the palate (*P*=0.024), and *Candida glabrata* on the denture (*P*=0.001).

There were no statistically significant differences in colony-forming unit (CFU) counts of *Candida* spp. on the dentures and palate (*P*=0.780; *P*=0.623), *Candida albicans* (*P*=0.505; *P*=0.881), *Candida tropicalis* (*P*=0.644; *P*=0.100), *Candida glabrata* (*P*=0.851; *P*=0.953), *Staphylococcus spp.* (*P*=0.689; *P*=0.728), and enterobacteria between the groups (Tables S3 and S4). However, the CFU count of mutans group streptococci in the dentures was higher in G3 (*P*=0.029). Nevertheless, no association was observed between this microorganism and SBP (*P*=0.339). A negative association between the CFU count of mutans group streptococci on the dentures with cardiovascular diseases (CVD) (*P*=0.006; r=-0.305) and the use of diuretics (*P*=0.005; r=-0.312) was noted (Table 2). Additionally, there was a positive association between the age of the participants and the CFU count of *Staphylococcus* spp. in the dentures (*P*=0.005, r=0.312), as illustrated in Tables S3 and S4.

In the assessment of diseases and medication declaration, the mean SBP was notably higher in the G3 (*P*=0.000). Among patients diagnosed with CVDs (n=5), 60% (n=3) were from G1, and 40% (n=2) were from G3. Furthermore, significant positive associations were found for the use of total antihypertensives (*P*=0.000; r=0.406), angiotensin II receptor blockers (*P*=0.019; r=0.263), and angiotensin-converting enzyme inhibitors (*P*=0.002; r=0.349), with a higher mean observed in groups using medications (G1 and G2) (*P*=0.000) (Table 4).

**Table 4.**
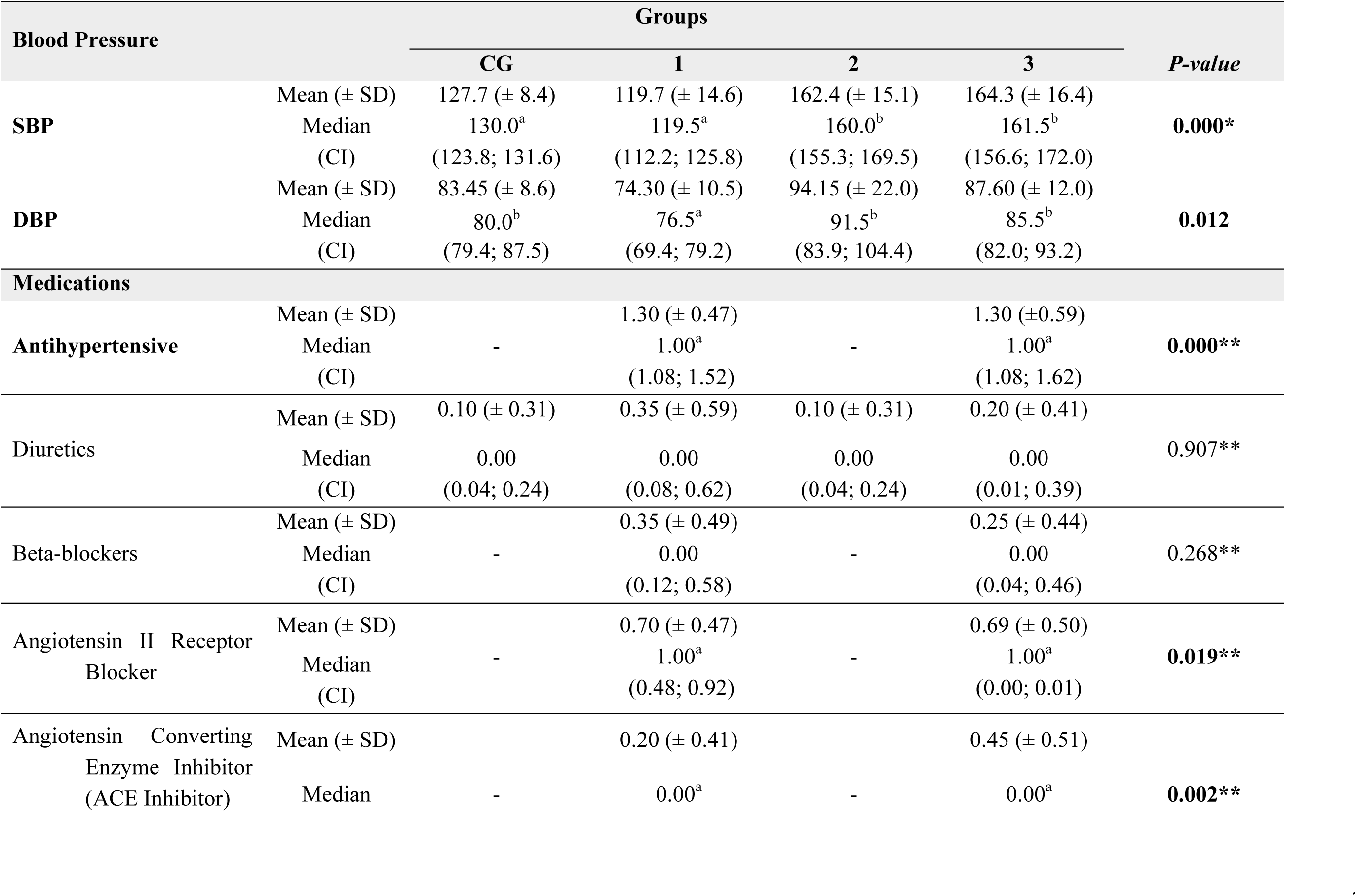
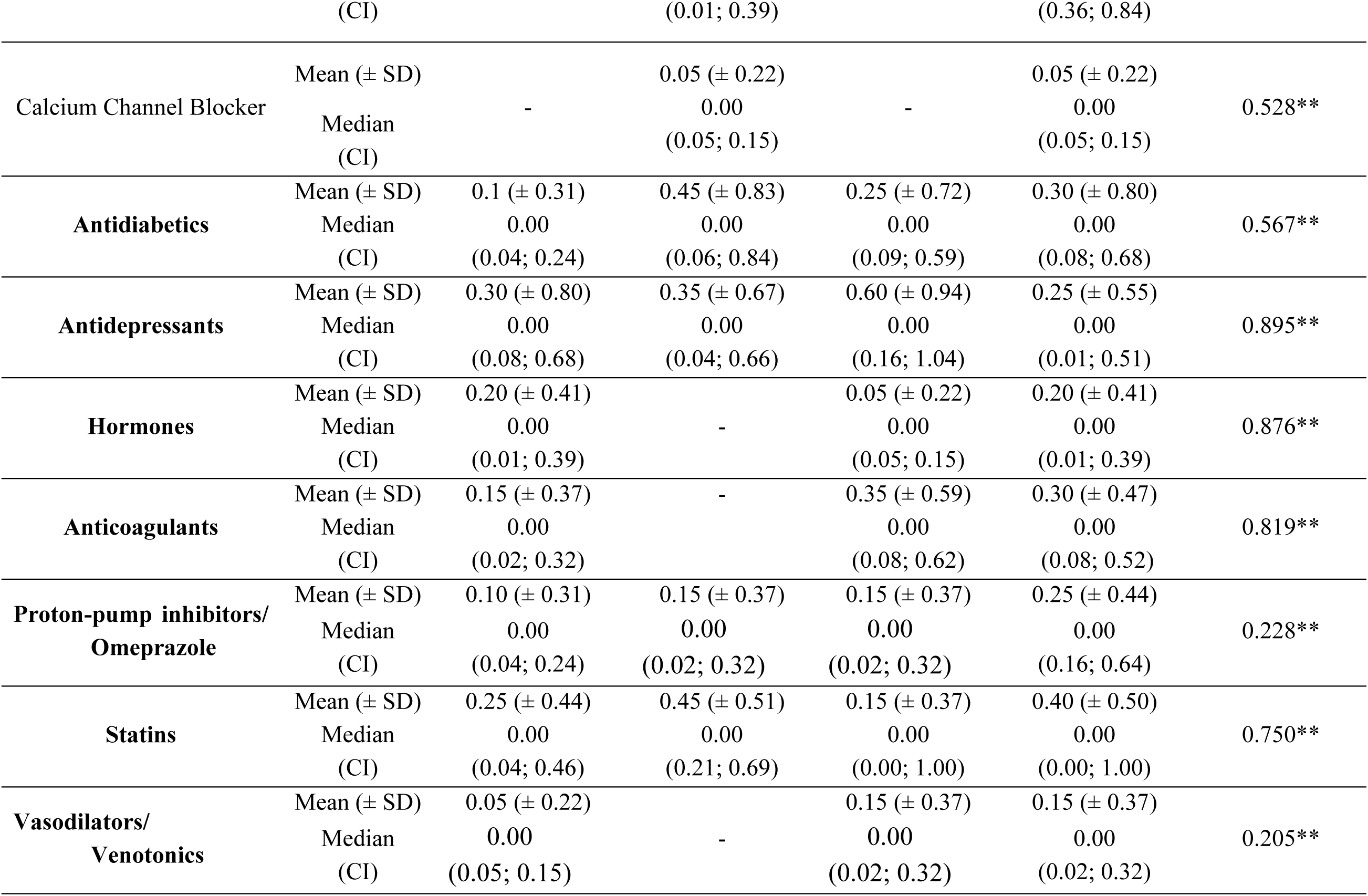
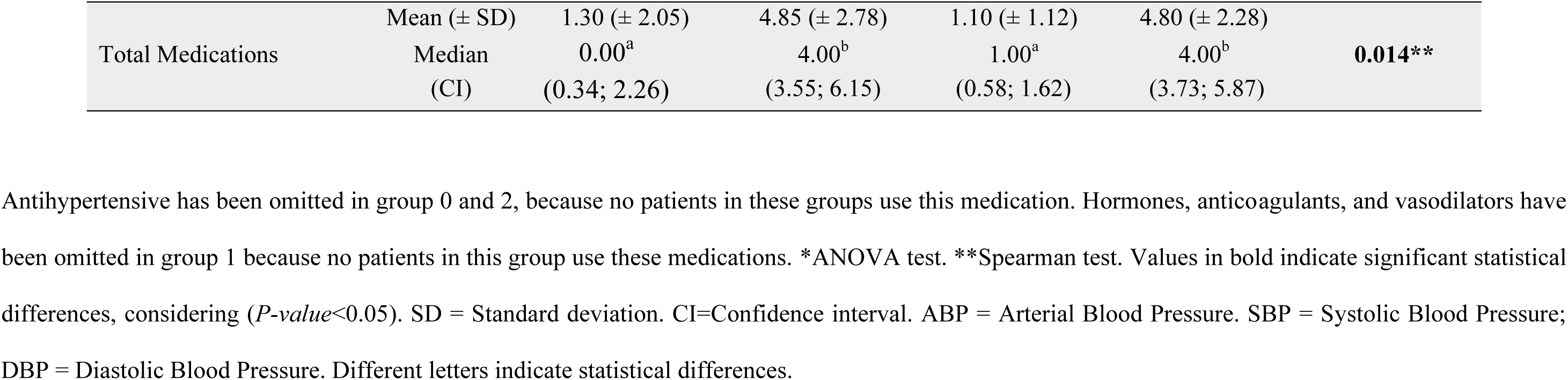
Survey of blood pressure and medications [Mean (SD); Median (CI)] of study participants.

## DISCUSSION

This case-control observational study investigated the microbial load present in the biofilm of dentures and the palate, as well as salivary flow and levels of inflammatory cytokines in edentulous individuals, users of maxillary dentures, normotensive individuals (group CG), and those affected due to hypertension (G1, G2, and G3). The results suggest that age and duration of edentulism may be associated with high systolic blood pressure. Furthermore, a possible association is observed between smoking habit and the expression of inflammatory cytokines, microbial load of mutans group streptococci, and the severity of cardiovascular diseases. Therefore, based on these findings, the null hypothesis was partially rejected.

Regarding sociodemographic characteristics, most participants were over the age of 60, female, married, living with their families, had completed primary education, and earned between 1 and 3 minimum wages in reais, consistent with findings from previous studies.^15–17,26,34^ Furthermore, elderly patients, irrespective of group allocation, exhibited a positive association with hypertension, aligning with results reported in prior research.^35–37^

In terms of behavioral habits, there is a tendency among CG participants to remove upper dentures at night (52.6%). Conversely, in G1, the majority reported occasionally removing both upper (65.0%) and lower (64.2%) dentures during the night, with no statistical difference in this habit between the groups. However, previous studies^38–41^ suggest that wearing dentures at night is linked to increased biofilm formation and a higher incidence of DS, factors that may contribute to the development of CVD.^42^ Dysbiosis of the oral microbiota and the presence of oral inflammatory conditions can trigger the production of inflammatory cytokines, according to observed in this study with TNF-α, leading to the release of C-reactive protein and the activation of a cytokine cascade, thereby promoting low-grade systemic inflammation.^43^

This study revealed a negative association between the number of cigarettes smoked daily and colony-forming unit (CFU) of mutans group streptococci on the dentures and a positive association with salivary levels of IL-4, IL-17, and IFN-γ. Smoking exerts a modulating effect on the microbial composition of various organs and systems in the human body, including the upper intestine, respiratory tract, and oral cavity.^19,44^ Numerous studies have indicated differences in the microbiota of smokers compared to non-smokers.^19,44^ For instance in a study of the Qatari population’s microbiome profile,^19^ found that non-smokers exhibit significantly greater diversity in their salivary microbiome compared to smokers, as evidenced by alpha diversity measures. Moreover, smoking activates inflammatory cells and elevates systemic levels of various inflammatory markers, such as C-reactive protein, fibrinogen, and IL-6,^44,45^ thus supporting the findings of our present study.

The results also suggest that there is a positive association between the number of cigarettes consumed daily and diabetic patients. This association can also be explained by the activation of inflammatory cytokines and changes in the oral microbiome triggered by smoking. The inflammatory response contributes to the occurrence of type 2 diabetes mellitus, causing insulin resistance, which can be intensified in the presence of hyperglycemia.^46^ Furthermore, changes in the oral and intestinal microbiome can affect the cardiometabolic profile and the host’s immune response.^47^

Moreover, a negative association was observed between the microbial load of mutans group streptococci on the dentures and CVDs, as well as the use of diuretics. Notably, patients in G1 reported a higher frequency of CVD diagnoses. Although this group comprises hypertensive patients showing a positive response to antihypertensives, it’s worth considering that G1 includes a larger proportion of smokers and ex-smokers. Given that daily cigarette consumption is linked to a reduction in the mutans group streptococci and is also associated with CVD, smoking could potentially act as a confounding factor for the negative association between this microorganism and CVD.

The negative association between mutans group streptococci and diuretics suggests a potential influence of diuretic use on the oral microbiome, especially in uncontrolled hypertensive patients (G3). Previous research has shown lower microbiota diversity in hypertensive patients,^48^ with evidence indicating alterations in microbial composition due to medication use. While this study cannot establish causality due to its observational nature and the possibility of medication interactions, it reinforces the need for further investigation into the relationship between medication use and oral microbiome changes,^50^ particularly in denture wearers, who are susceptible to potential shifts in the oral microbiome composition.

Edentulism has consistently been linked to hypertension in various studies.^6–11^ Individuals with hypertension tend to exhibit a higher prevalence of missing teeth compared to those without the condition.^26^ Previous research also indicates a greater incidence of coronary heart disease, stroke, and elevated systolic blood pressure among individuals with extensive tooth loss. ^24,51^ In this study, while a direct relationship between tooth loss and systolic blood pressure could not be established as all patients were edentulous, an examination of the duration of edentulism revealed a significantly longer period in the G3, with a positive association observed between this duration and blood pressure. It is crucial to highlight that the group with the highest prevalence of smokers and ex-smokers (G1) exhibits a positive relationship with antihypertensive medications, while the group with the poorest response to medications (G3), despite having fewer smoking habits, demonstrates higher levels of hypertension. This suggests that there may be an immunological factor influencing these responses to hypertension, rendering G3 more predisposed to an unfavorable systemic condition. In this context, the period of edentulism could represent an unconventional factor contributing to this systemic condition.

Furthermore, during the analysis of patients who lacked functional rehabilitation, specifically those with only upper dentures, higher levels of IL-2 cytokines were observed in hypertensive patients not controlled with medication (G3). This finding may be explained by the longer time of edentulism observed in this group and its correlation with reduced masticatory function and altered food choices. Research indicates that tooth loss can lead to detrimental changes in dietary habits, including decreased intake of vitamin C, fiber, fruits, and vegetables, along with increased consumption of high-fat foods such as processed and fatty foods.^6,52,53^ Elevated body mass index (BMI) resulting from dietary changes can lead to adipose tissue dysfunction, characterized by increased expression of cytokines that promote vascular permeability and recruit immune system cells, thereby triggering a state of low-grade chronic inflammation.^42,54^

In this context, a previous study^55^ discovered that women with fewer teeth tend to decrease their consumption of fruits and vegetables, potentially elevating the risk of CVD. Additionally, tooth loss has been linked to a substantial risk of CVD and incidences of stroke. For each additional loss of two teeth, there was a 3% rise in the risk of both coronary heart disease and stroke.^56^

Furthermore, the concentration of mutans group streptococci in the oral cavity may also be influenced by diet. Frequent exposure to dietary carbohydrates can create a dysbiotic oral environment, leading to increased acid concentrations in the oral cavity. This environment may favor the predominance of acid-tolerant species like *Streptococcus mutans* over less acid-tolerant commensal bacteria in dental plaque. Moreover, the acidogenic and aciduric properties of this environment fostered by *S. mutans* can encourage the growth of yeasts, such as *Candida* spp.^57,58^ However, our study did not assess the patient’s body mass index or conduct nutritional monitoring, which could be addressed in future research endeavors.

It was observed that patients with a more severe degree of DS had higher CFU counts for *Candida* spp. both on the palate and on the denture, *Candida albicans* on the denture, *Candida tropicalis* on the palate and *Candida glabrata* on the denture. Poor hygiene is one of the main causes of DS, as it contributes to the high prevalence of *Candida* spp. and other microorganisms that complement each other.^16,32,59^

Although this study did not observe a significant difference between the groups concerning DS and did not establish an association of this oral condition with hypertension and CVD, previous research suggests that oral bacteria implicated in the etiopathogenesis of chronic periodontitis and DS can trigger a moderate systemic immunological and inflammatory response. These findings indicate an elevation in serum levels of various cytokines and inflammatory markers, which are prominently produced in pathological oral tissues and strongly linked to the pathogenesis of certain CVD.^60,61^

In addition, elderly patients exhibited higher CFU of *Staphylococcus* spp. on the dentures. Recent studies suggest a correlation between the aging process and alterations in the microbiota.^18,62,63^ Senescence has been linked to reduced diversity in the oral microbiome and an increase in pathogenic colony counts in saliva, including periodontopathogens such as *Aggregatibacter actinomycetemcomitans*, *Porphyromonas gingivalis*, *Treponema denticola*, and *Tannerella forsythia*.^19^ Furthermore, research indicates that strains of *Staphylococcus aureus* produce an inhibitory compound akin to bacteriocin, a thermostable molecule with a protein half-life, capable of inhibiting gram-positive bacteria. Similarly, the species *S. mutans* can inhibit the growth of other bacteria like *Treponema denticola* through the production of acids and reduction of pH.^64^

Furthermore, *Staphylococcus* spp. it was also associated with the presence of dyslipidemia, mainly with cholesterol. This data corroborates previous studies that indicated a reduction in total cholesterol in models infected by *Staphylococcus aureus*. This reduction indicates that sterol metabolism may be altered during bacterial intestinal infection, mainly due to the disorganization of the epithelial surface membrane, causing symptoms such as intestinal irritation.^65,66^

Although patients in the CG reported more xerostomia, no significant differences were observed in salivary flow levels between groups. Oral microorganisms typically thrive in areas with reduced salivary flow, such as interdental spaces, gingival crevices, and denture- fitting surfaces.^67^ However, since all patients in our study were elderly denture wearers, this might explain the absence of significant differences in salivary flow levels between the groups. Conversely, we found a negative association between the severity of DS and decreased salivary flow, consistent with findings from other studies.^15,16^ Additionally, male individuals exhibited lower salivary flow compared to women, contrary to findings in other studies,^68,69^ suggesting a potential association with external factors such as medication use and behavioral habits.

This study has certain limitations as it is an observational study and cannot establish a causal correlation between the microbiota of individuals using complete dentures and CVD. Moreover, randomized controlled studies encompassing younger and dentate patients are essential to elucidate the relationship among age, edentulism, microbiota, and hypertension. Additionally, the oral microbiota exhibits considerable diversity, and not all microorganisms are cultivable, rendering the cultivation technique on selective media ineffective in detecting specific pathogenic species. Studies employing more advanced techniques such as genetic sequencing are necessary to enhance the accuracy and reliability of this analysis.

## PERSPECTIVES

It is suggested that both age and duration of edentulism may contribute to hypertension, while smoking habits may modulate the salivary microbiota of the mutans group streptococci, triggering a cascade of interleukins and exerting a greater influence, particularly on diabetic or CVD patients. Additionally, non-functional dental rehabilitations exhibit a more pronounced association with hypertensive patients not controlled by medication, as reflected in elevated systolic blood pressure and IL-2 levels.

In conclusion, the interaction between oral health and systemic conditions like hypertension involves a complex interplay of factors including age, duration of edentulism, smoking habits, salivary microbiota, and the quality of dental rehabilitations. Understanding these relationships underscores the importance of comprehensive healthcare approaches that address both oral and systemic health factors for effective disease management and prevention.

## Data Availability

All data referenced in this manuscript may be made available upon request from the authors.

## NOVELTY AND RELEVANCE

### What Is New?

To our knowledge, this study proposal represents the first attempt to concurrently investigate the microbial load of the dentures and palate, salivary flow, and salivary levels of inflammatory cytokines in normotensive and hypertensive edentulous patients utilizing complete dentures. Furthermore, information regarding the duration of edentulism and denture usage can contribute as essential parameters for medical clinics and cardiologists, particularly when considering patients with multiple risk factor associations.

### What Is Relevant?

The study reveals intriguing findings regarding how behavioral habits, particularly smoking, can impact the oral microbiota and be linked to inflammatory cytokines. Furthermore, there is a discussion on how aspects of dental history, such as the duration of edentulism, may be associated with hypertension, and the quality of oral rehabilitation is highlighted as a factor that can influence SBP and IL-2 levels.

### Clinical/Pathophysiological Implications?

Clinical findings offer crucial insights into the oral health of both normotensive and hypertensive patients, highlighting the significance of multidisciplinary care in addressing atypical factors among hypertensive individuals. Understanding mechanisms beyond these conditions can aid in the prevention and reduction of exacerbation of oral and systemic diseases within primary care, potentially leading to significant impacts on public health costs.

## AUTHORS CONTRIBUITION

Pizziolo PG and Clemente LM contributed to the biofilm collection, instructed participants to answer specific questionnaires, laboratory processing, data tabulation and analysis, and wrote the article. Oliveira VC classified the microorganisms’ species. Ribeiro AB^2^ quantified the concentration of salivary levels of cytokines. Macedo AP contributed to statistical analyses and interpretation of data. Salgado HC, Fazan-Junior R, Watanabe E, and Silva-Lovato CH critically revised the article for important intellectual content. Ribeiro AB contributed to the conception, design, and administration of the study.

## Nonstandard Abbreviations and Acronyms

CVD: cardiovascular disease
SF: salivary flow
SBP: systolic blood pressure
DBP: diastolic blood pressure
CFU: colony-forming unit
DS: denture stomatitis
PBS: phosphate buffered saline
ACE Inhibitor: angiotensin converting enzyme inhibitor
SD: standard deviation
CI: confidence interval

